# Identifying COVID-19 phenotypes using cluster analysis and assessing their clinical outcomes

**DOI:** 10.1101/2022.05.27.22275708

**Authors:** Eric Yamga, Louis Mullie, Madeleine Durand, Alexandre Cadrin-Chenevert, An Tang, Emmanuel Montagnon, Carl Chartrand-Lefebvre, Michaël Chassé

## Abstract

Multiple clinical phenotypes have been proposed for COVID-19, but few have stemmed from data-driven methods. We aimed to identify distinct phenotypes in patients admitted with COVID-19 using cluster analysis, and compare their respective characteristics and clinical outcomes.

We analyzed the data from 547 patients hospitalized with COVID-19 in a Canadian academic hospital from January 1, 2020, to January 30, 2021. We compared four clustering algorithms: K-means, PAM (partition around medoids), divisive and agglomerative hierarchical clustering. We used imaging data and 34 clinical variables collected within the first 24 hours of admission to train our algorithm. We then conducted survival analysis to compare clinical outcomes across phenotypes and trained a classification and regression tree (CART) to facilitate phenotype interpretation and phenotype assignment.

We identified three clinical phenotypes, with 61 patients (17%) in Cluster 1, 221 patients (40%) in Cluster 2 and 235 (43%) in Cluster 3. Cluster 2 and Cluster 3 were both characterized by a low-risk respiratory and inflammatory profile, but differed in terms of demographics. Compared with Cluster 3, Cluster 2 comprised older patients with more comorbidities. Cluster 1 represented the group with the most severe clinical presentation, as inferred by the highest rate of hypoxemia and the highest radiological burden. Mortality, mechanical ventilation and ICU admission risk were all significantly different across phenotypes.

We conducted a phenotypic analysis of adult inpatients with COVID-19 and identified three distinct phenotypes associated with different clinical outcomes. Further research is needed to determine how to properly incorporate those phenotypes in the management of patients with COVID-19.

## Introduction

Patients affected with coronavirus disease 2019 (COVID-19) have shown significant clinical heterogeneity and variability in disease trajectory (1). Clinical phenotypes are homogeneous subgroups of a disease presenting distinct clinical features (2). Well-established phenotypes are potentially useful at the bedside for appropriately classifying patients into meaningful categories, predicting disease course, and personalizing treatments.

Since the first case description of COVID-19, various phenotypes have emerged, each using various layers of clinical information. Two phenotypes have been described based on lung mechanics and radiological findings (3,4), others have focused on disease complications—as such, a hypercoagulable phenotype has been observed, prompting recommendations for intensified antithrombotic therapy (5–7).

The majority of those phenotypes failed to fully describe the complexity of the disease as they focused on characterizing one dimension of the clinical presentation. Being mostly derived from clinical observation, or based on outcomes that can only occur in the future is however potentially less relevant in a prospective and real-world context. Thus, the reliability and the methodology of those first phenotyping efforts have been put into question (8,9). Increasing amounts of medical data allow conducting phenotypic analyses using data-driven techniques. Those methods are hypothesis-agnostic and solely rely on the assumption that clinical patterns lie within the data (10). Clustering is an unsupervised machine learning method used to identify homogeneous groups within a heterogeneous dataset. This method has been used to describe clinical phenotypes in other diseases such as asthma (11,12), COPD (13) and sepsis (14).

The primary objective of this study was to identify COVID-19 cluster of phenotypes at patient presentation using multimodal real-world clinical data and medical imaging data (15). Our secondary objectives were to assess the association of those phenotypes with three clinical outcomes: mechanical ventilation (MV), intensive care unit (ICU) admission and hospital mortality. Finally, we created a simple decision tree classifier allowing the interpretation and assignment of patients to one of the identified phenotypes.

## Methods

### Data sources

We used real-world data extracted from clinical source system comprising relevant clinical information from all COVID-19-related hospitalizations at the Centre for the Integration and Analysis of Medical Data (CITADEL) of the Centre Hospitalier de l’Université de Montréal (CHUM), a Canadian academic quaternary center. The analytical dataset contains de-identified data for over 1,100 patients hospitalized with COVID-19, including demographics, comorbidities, laboratory results, vital signs, drugs, medical procedures, frontal chest radiographs (CXR) and clinical outcomes. The raw data was managed using SQLite 3, and further data processing was conducted using Python version 3.7 and R version 4.0.3. We provided additional details regarding initial data processing. (see **S1 text, supplementary methods**).

### Study population

We included all unique adult hospitalizations (≥ 18 years of age) for COVID-19 from January 1, 2020, to January 30, 2021, for which a chest X-ray was available within 24 hours of admission. A COVID-19 hospitalization episode was defined as a hospitalization within seven days of a positive SARS-CoV-2 PCR result. The Institutional Review Board of the CHUM (Centre Hospitalier de l’Université de Montréal) approved the study and informed consent was waived because of its low risk and retrospective nature.

### Imaging Data Processing

Imaging data were obtained in the DICOM format, and frontal CXR (posteroanterior and anteroposterior) were processed, discarding lateral CXR. Lung opacities observed on CXR were manually annotated with bounding boxes by a board-certified radiologist using a bounding box annotation software (16). This annotation method is recognized by the Radiological Society of North America (RSNA) and is the annotation methodology of choice for all deep learning challenges involving image detection (17,18). Bounding boxes are rectangular or squared delimitations of the opacities found on a given chest radiograph reported with a scaled width and a scaled length. Hence, from the manual annotation, we derived the number of opacities and the total size of opacities as a relative percentage of the total surface of the image.

### Variables Selection and Feature Engineering

From the analytical dataset, we extracted 160 candidate variables. We provided the list of those variables in the supplementary material (see **S1 Table**). For each of those variables, we exclusively used the first recorded value within the first 24 hours of admission. We then excluded 56 variables for which more than 25% of observations were missing. The remaining missing variables were imputed using all available features except clinical outcomes (ICU admission, mechanical ventilation and death). We used classification and regression tree (CART) single mean imputation, a robust method against outliers, multicollinearity and skewed distributions that has the benefit of being simple to implement in a real-world setting (19). We acknowledge that other imputation algorithms such as Expectation Maximization (EM) and Multiple Imputation (MI) have shown superior performance but the additional computational cost makes those approaches difficult to implement in a real-world setting (20).

We then computed three more variables that have recently been associated with COVID-19 mortality: neutrophil-to-lymphocyte ratio (NLR) (21), the ratio of peripheral arterial oxygen saturation to the inspired fraction of oxygen (SpO_2_/FiO_2_) (22), and the shock index (heart rate/systolic blood pressure) (23). We also computed the Medicines Comorbidity Index (MCI), a metric to assess multimorbidity that has shown similar epidemiological value to the Charlson Comorbidity Index (CCI) (24). We relied on the MCI instead of the CCI because comorbidities were not systematically recorded in our database, but home medications were (see **S1 Table**). MCI was computed at the time of study enrollment only using data that was available on presentation to mimic real-world setting. We summarized the final set of variables included for analysis with their respective subdomains of interest in **Table 1**.

**Table 1.** Final set of variables and respective subdomains.

| Subdomain | Clinical variable |
| --- | --- |
| Demographic | Age, sex, comorbidities (MCI) |
| Hemodynamic | SBP, DBP, HR, Shock index (HR/SBP), Anion Gap |
| Respiratory | SpO<sub>2</sub>, SpO<sub>2</sub>/FiO<sub>2</sub> |
| Imaging | Opacities number, Opacities size |
| Hematologic | Neutrophils, Lymphocytes, NLR |
| Inflammatory/Thrombotic | MPV, Temperature |
| Renal | Creatinine, Sodium, Potassium, Bicarbonate |

Despite receiving little attention, feature engineering in clustering efforts has shown to increase learning performance through dimensionality reduction (25). In the healthcare setting, aggregated variables such as the one included in our algorithm also enhances the stability of a learning model’s performance over time (26). In addition to the statistical advantages mentioned, feature engineering also allows for better interpretability, as aggregated variables are often clearer and more intuitive. Pre-processed variables do not impede the hypothesis-agnostic premise of unsupervised machine learning, as all these variables are directly derived from the features of the original dataset and no preferential weighting was conducted.

### Data Preparation and Cluster Analysis

Before applying clustering algorithms, we adequately processed our dataset, log-transforming skewed continuous variables (skewness > 0.5) and excluding highly correlated variables (correlation > 0.8). The then obtained principal components via factor analysis of mixed data (FAMD) (27) for all our observations. The excluded variables based on high correlation were the total white blood cell count (WBC), the Fraction of Inspired Oxygen (FiO2) and the heart rate (HR). Those three variables were respectively highly correlated with neutrophils, the SpO_2_/FiO_2_ ratio and the shock index. We provided additional details regarding the data preparation in the supplementary material (see **S1 text**).

We compared four clustering algorithms: K-means, PAM (partition around medoids), divisive and agglomerative hierarchical clustering. We used three internal validation metrics (connectivity, Dunn index, the average silhouette width) and four stability measures (average proportion of non-overlap (APN), the average distance (AD), the average distance between means (ADM), and the figure of merit (FOM)) to compare the algorithms. The optimal algorithm and the optimal number of clusters were then determined by rank aggregation of the ranked lists of each validation metric. We used the optCluster package, an R package facilitating the execution of the aforementioned analyses (28).

### Phenotypes Robustness: sensitivity analyses

We conducted a sensitivity analysis to assess whether the removal of imaging data altered the performance of the clustering algorithm. We first compared the clustering results given these two scenarios using the average silhouette width and the adjusted rand index (29), a measure of the similarity between two data clustering. We then assessed the number of patients that underwent phenotypic reclassification before and after the removal of imaging data in the clustering algorithm. In other words, we analyzed the characteristics of patients for which the cluster assignment differed after the removal of imaging data.

## Statistical Analysis

### Clusters Interpretation

We assessed the difference in the distribution of features across identified clusters using the Chi-Square test for categorical variables, analysis of variance (ANOVA) and Kruskal–Wallis test for normally distributed and non-normally distributed continuous variables respectively.

To facilitate the interpretation and clinical usability of the obtained clusters, we trained a simple decision tree using the CART with the clusters as predicted outcomes (30). We also determined the most critical variables to distinguish the clusters by conducting a variable importance analysis (VIA) (31). Details regarding VIA were provided in the supplementary material (see **S1 text, S1 Figure**).

### Clinical Outcomes Evaluation

We conducted survival analysis using Kaplan-Meier method to compare clinical outcomes according to clusters. We assessed three clinical outcomes: 7-day ICU admission, 7-day mechanical ventilation and 30-day mortality. To reduce confounding bias, we specifically restricted clinical outcomes analysis to patients eligible for MV and ICU admission according to their Physician Orders for Life-Sustaining Treatment (POLST) form.

## Results

### Study Population

In total, 1,125 unique COVID-19 hospitalizations were screened. A total of 559 patients were excluded after removing readmissions (*n* = 36), patients without a CXR within 24 hours of admission (*n* = 523), and patients for which clinical data was missing (*n =* 19), leaving 547 patients for the cluster analysis (see **Fig 1**). We provided details regarding the characteristics of our study cohort (see **Table 2**). Our population was similar to other cohorts of patients hospitalized with COVID-19 in North America during the first two waves, with a mean age of 69 years old, a relatively similar proportion of men and women, and an in-hospital mortality rate of 20% (32).

**Table 2.** Baseline characteristics of the study population.

| Characteristics | Count (%) or Mean(SD) or Median [IQR] |
| --- | --- |
| N | 547 |
| Age (years) | 66.56 (17.94) |
| Sex (male, %) | 313 (57.2) |
| **Laboratory results** |  |
| Hemoglobin | 127.23 (20.69) |
| Platelet | 207.00 [157.00, 271.50] |
| WBC | 6.80 [5.30,9.85] |
| Neutrophil count | 5.01 [3.60,7.60] |
| Lymphocyte count | 0.95 [0.62,1.35] |
| Monocytes count | 0.63 (0.39) |
| Basophils count | 0.01 (0.03) |
| Eosinophils count | 0.05 (0.12) |
| VPM | 9.89 (1.33) |
| VGM | 89.90 [85.90,93.85] |
| NLR | 5.25 [3.19,10.00] |
| Sodium | 137.59 (5.05) |
| Potassium | 4.00 (0.53) |
| Bicarbonate | 24.95 (3.76) |
| Anion gap | 11.05 (3.69) |
| Creatinine (μmol/L) | 78.00 [63.00,105.00] |
| **Vital Signs** |  |
| FiO<sub>2</sub> | 21.00 [21.00,28.00] |
| SpO<sub>2</sub> | 95.00 [94.00,97.00] |
| SpO<sub>2</sub>/FiO<sub>2</sub> | 447.62 [339.29,461.90] |
| Temperature | 36.98 (0.51) |
| Systolic Blood Pressure | 130.00 [116.00,144.50] |
| Diastolic Blood Pressure | 75.00 [68.00, 82.00] |
| Heart Rate | 92.2 (20.2) |
| Shock Index | 0.69 [0.58,0.82] |
| Respiratory Rate | 20.00 [20.00,24.00] |
| **Home Medication** |  |
| Medicines Comorbidity Index | 2.77 (2.04) |
| Anticholesterolemic agents | 183 (33.5) |
| **Antihypertensive agents** | 241 (44.1) |
| **Bronchodilator agents** | 178 (32.5) |
| **Diuretics** | 137 (25.0) |
| **Factor Xa Inhibitors** | 50 ( 9.1) |
| **Hypoglycemic agents** | 209 (38.2) |
| **Platelet aggregation inhibitors** | 150 (27.4) |
| **Imaging Data** |  |
| **Opacities Numbers (%)** |  |
| **0** | 141 (25.8) |
| **1** | 89 (16.3) |
| **2** | 279 (51.0) |
| **3** | 37 ( 6.8) |
| **4** | 1 ( 0.2) |
| **Opacities Size (surface area %)** | 7 [0.0,15] |
| **Clinical outcomes** |  |
| **Length of Stay (days)** | 8.29 [3.48, 18.20] |
| **Mechanical Ventilation** | 48 ( 8.8) |
| **Wave (1st)** | 295 (53.9) |
| **ICU admission** | 132 (24.1) |
| **Death** | 113 (20.7) |

Count (%) or Mean (SD) or Median [IQR]

**Figure 1.**
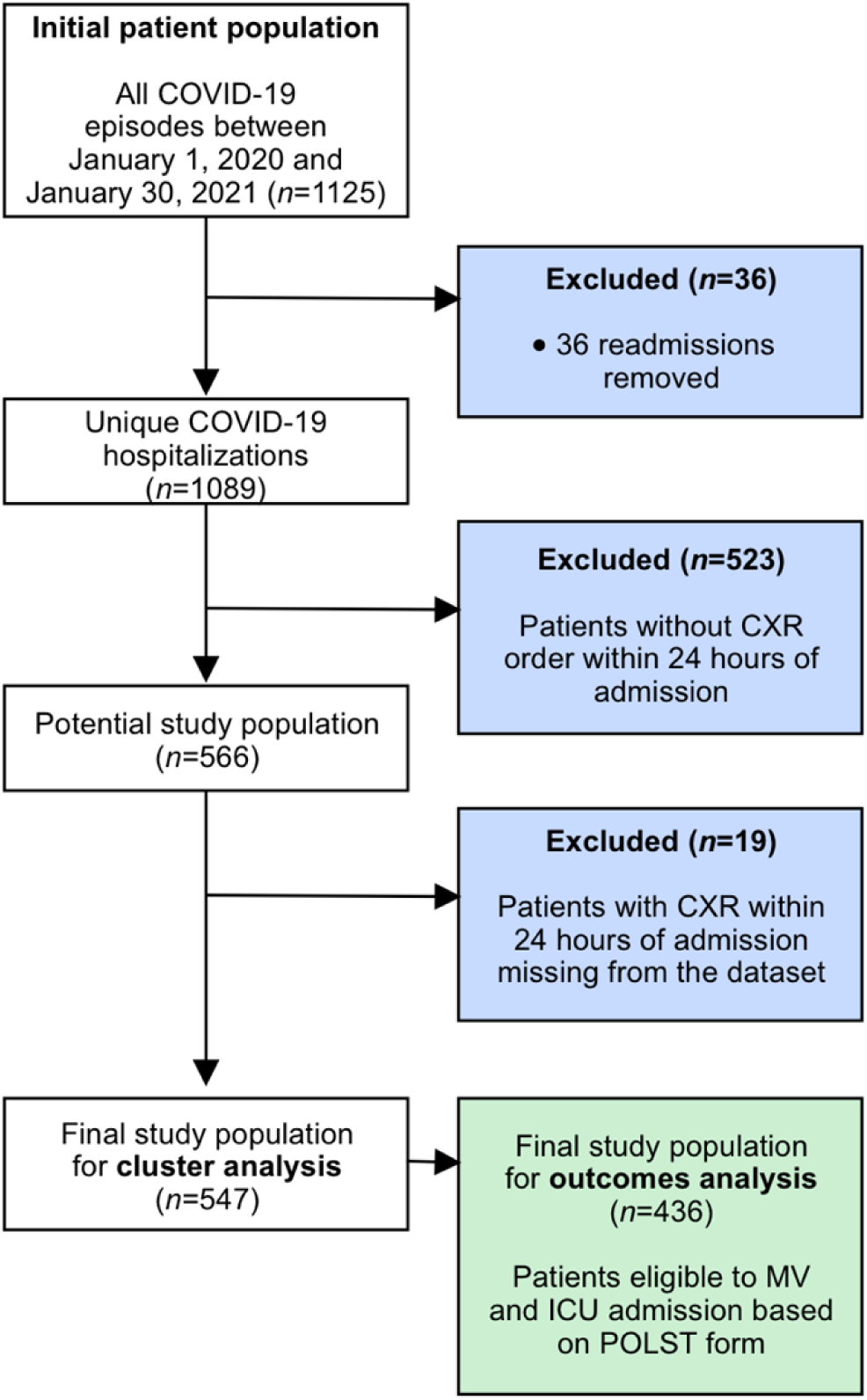
Study Inclusion and Exclusion Criteria

### Clinical Characteristics of Phenotypes

Agglomerative hierarchical clustering was deemed the most robust clustering algorithm for our dataset. The optimal number of K clusters was K = 3 (see **Fig 2**), yielding the highest clustering performance as exhibited by the rank aggregation of seven internal validation measures (see **S2 Figure, S2 Table**).

**Figure 2.**
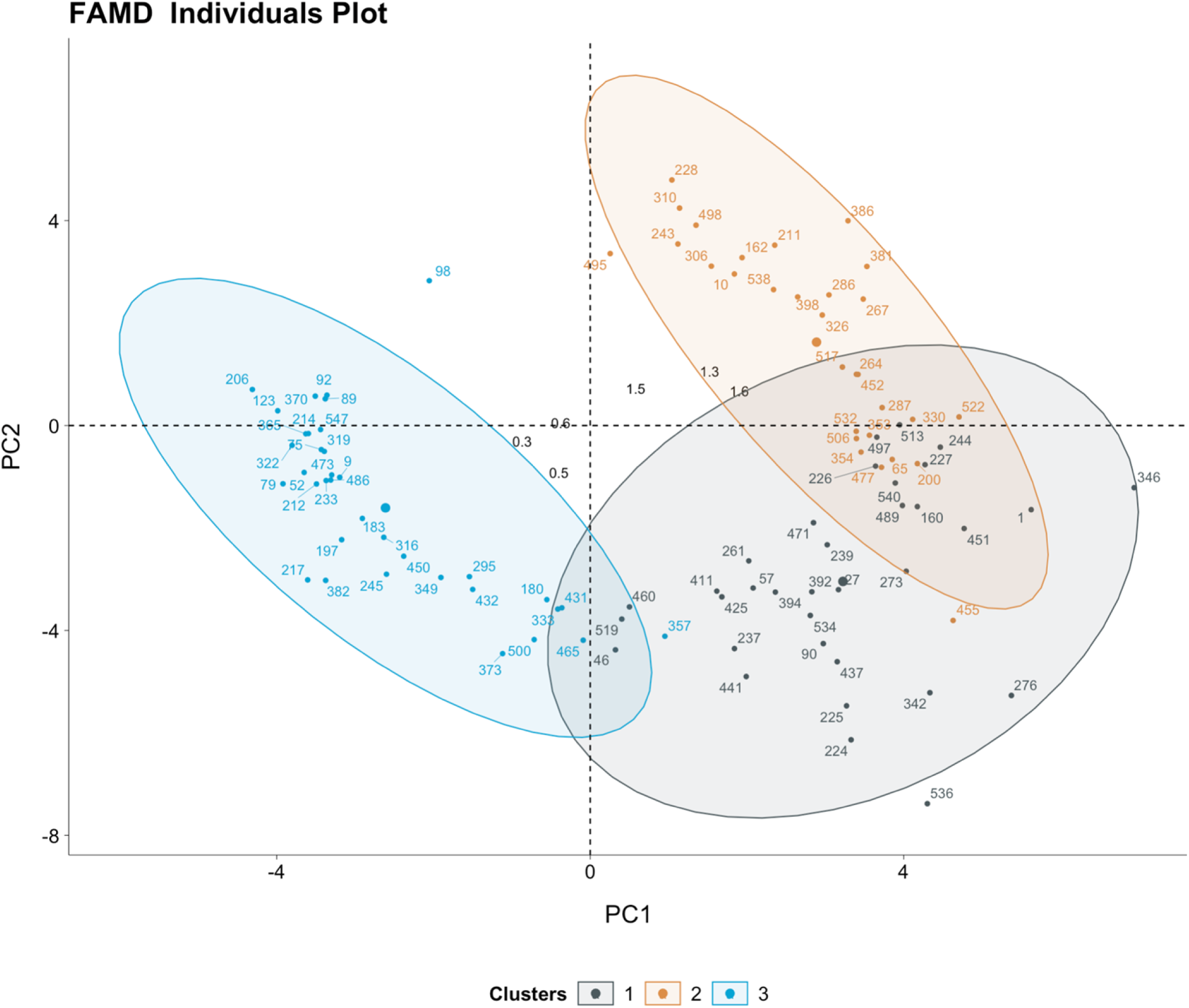
Clusters visualization on factor analysis of mixed data principal components plot

We summarized the characteristics regarding those clusters in **Table 3**. As determined through the VIA, the most important variables for discriminating clusters were age, the MCI, the SpO_2_/FiO_2_ ratio, opacities size, the neutrocyte-lymphocyte ratio, creatinine and the shock index (see **Figures 3A-3B**).

**Table 3.** Clinical characteristics stratified by clusters.

|  | Cluster 1 | Cluster 2 | Cluster 3 | p-value |
| --- | --- | --- | --- | --- |
| N (%) | 61 (11%) | 211 (39%) | 275 (50%) |  |
| Age (years) | 66.18 (15.01) | 75.46 (12.54) | 59.82 (19.10) | <0.001 |
| Sex (male) | 26 (42.6) | 123 (58.3) | 164 (59.6) | 0.695 |
| **Laboratory results** |  |  |  |  |
| Hemoglobin | 117.69 (21.57) | 125.55 (20.83) | 130.43 (20.35) | <0.001 |
| Platelet | 244.00 [187.00, 333.00] | 195.00 [148.50, 259.00] | 207.00 [160.50, 269.00] | 0.004 |
| WBC<sup>†</sup> | 9.80 [6.90, 13.60] | 6.70 [5.35, 9.80] | 6.60 [4.90, 9.25] | <0.001 |
| Neutrophil count | 8.70 [5.82, 12.63] | 5.03 [3.76, 7.46] | 4.63 [3.25, 6.92] | <0.001 |
| Lymphocyte count | 0.60 [0.35, 0.98] | 0.90 [0.60, 1.30] | 1.06 [0.76, 1.54] | <0.001 |
| Monocytes count | 0.51 (0.31) | 0.68 (0.42) | 0.61 (0.37) | 0.006 |
| Basophils count | 0.01 (0.03) | 0.02 (0.03) | 0.01 (0.02) | 0.358 |
| Eosinophils count | 0.02 (0.04) | 0.06 (0.14) | 0.04 (0.09) | 0.010 |
| NLR | 13.53 [9.37, 25.22] | 6.00 [3.45, 10.20] | 4.36 [2.66, 7.08] | <0.001 |
| Sodium | 137.92 (5.56) | 138.08 (5.46) | 137.47 (4.58) | 0.764 |
| Potassium | 4.24 (0.68) | 4.08 (0.54) | 3.91 (0.45) | 0.013 |
| Bicarbonate | 22.78 (4.85) | 25.23 (3.76) | 25.20 (3.02) | <0.001 |
| Anion Gap | 13.22 (4.25) | 10.67 (3.64) | 10.87 (3.43) | <0.001 |
| Creatinine (μmol/L) | 84.00 [62.00, 140.00] | 91.00 [71.00, 126.00] | 72.00 [58.00, 87.00] | 0.002 |
| **Vital Signs** |  |  |  |  |
| FiO<sub>2</sub><sup>†</sup> | 80.00 [40.00, 100.00] | 21.00 [21.00, 24.50] | 21.00 [21.00, 21.00] | <0.001 |
| SpO<sub>2</sub> | 92.00 [88.00, 95.00] | 95.00 [94.00, 97.00] | 96.00 [94.00, 98.00] | <0.001 |
| SpO<sub>2</sub>/FiO<sub>2</sub> | 118.75 [92.00, 225.00] | 447.62 [368.75, 457.14] | 457.14 [442.86, 466.67] | <0.001 |
| Temperature | 36.89 (0.36) | 36.96 (0.52) | 37.00 (0.54) | 0.327 |
| Systolic Blood Pressure | 125.00 [116.00, 153.00] | 133.00 [120.00, 149.50] | 125.00 [114.00, 140.00] | <0.001 |
| Diastolic Blood Pressure | 75.00 (10.67) | 73.57 (12.31) | 76.34 (13.06) | 0.054 |
| Heart Rate<sup>†</sup> | 91.77 (18.05) | 89.61 (17.55) | 94.34 (22.33) | 0.037 |
| Shock Index | 0.72 [0.54, 0.86] | 0.66 [0.58, 0.76] | 0.73 [0.61, 0.86] | <0.001 |
| Respiratory Rate | 26.00 [22.00, 32.00] | 20.00 [18.00, 24.00] | 20.00 [18.50, 23.00] | <0.001 |
| **Home Medication** |  |  |  |  |
| **Medicines Comorbidity Index (MCI)** | 3.34 (1.89) | 4.30 (1.65) | 1.47 (1.38) | <0.001 |
| **Anticholesterolic agents** | 20 (32.8) | 133 (63.0) | 29 (10.5) | <0.001 |
| **Antihypertensive agents** | 22 (36.1) | 145 (68.7) | 69 (25.1) | <0.001 |
| **Bronchodilator agents** | 20 (32.8) | 79 (37.4) | 74 (26.9) | 0.046 |
| **Diuretics** | 28 (45.9) | 77 (36.5) | 30 (10.9) | <0.001 |
| **Factor Xa Inhibitors** | 1 ( 1.6) | 42 (19.9) | 5 ( 1.8) | <0.001 |
| **Hypoglycemic agents** | 40 (65.6) | 120 (56.9) | 50 (18.2) | <0.001 |
| **Platelet aggregation inhibitors** | 13 (21.3) | 113 (53.6) | 24 ( 8.7) | <0.001 |
| **Imaging Data** |  |  |  |  |
| **Opacities Numbers (%)** |  |  |  | <0.001 |
| **0** | 1 ( 1.6) | 34 (16.1) | 106 (38.5) |  |
| **1** | 4 ( 6.6) | 47 (22.3) | 38 (13.8) |  |
| **2** | 54 (88.5) | 106 (50.2) | 119 (43.3) |  |
| **3** | 2 ( 3.3) | 23 (10.9) | 12 ( 4.4) |  |
| **4** | 0 ( 0.0) | 1 ( 0.5) | 0 ( 0.0) |  |
| **Opacities Size (surface area %)** | 20 (8) | 9 (7) | 7 (7) | <0.001 |
| **Clinical outcomes** |  |  |  |  |
| **Length of stay (days)** | 14.99 [6.00, 34.42] | 9.95 [4.61, 20.04] | 5.04 [1.32, 11.51] | <0.00 |
| **Mechanical ventilation** | 24 (39.3) | 16 ( 7.6) | 8 ( 2.9) | <0.001 |
| **ICU admission** | 43 (70.5) | 46 (21.8) | 43 (15.6) | <0.001 |
| **Death** | 26 (42.6) | 51 (24.2) | 36 (13.1) | <0.001 |
| **Other** |  |  |  |  |
| **Wave (1st)** | 38 (62.3) | 122 (57.8) | 135 (49.1) | 0.061 |

Count (%) or Mean (SD) or Median [IQR]
† Those variables were excluded from the clustering algorithm effort because of the presence of other highly correlated variables

**Figure 3A.**
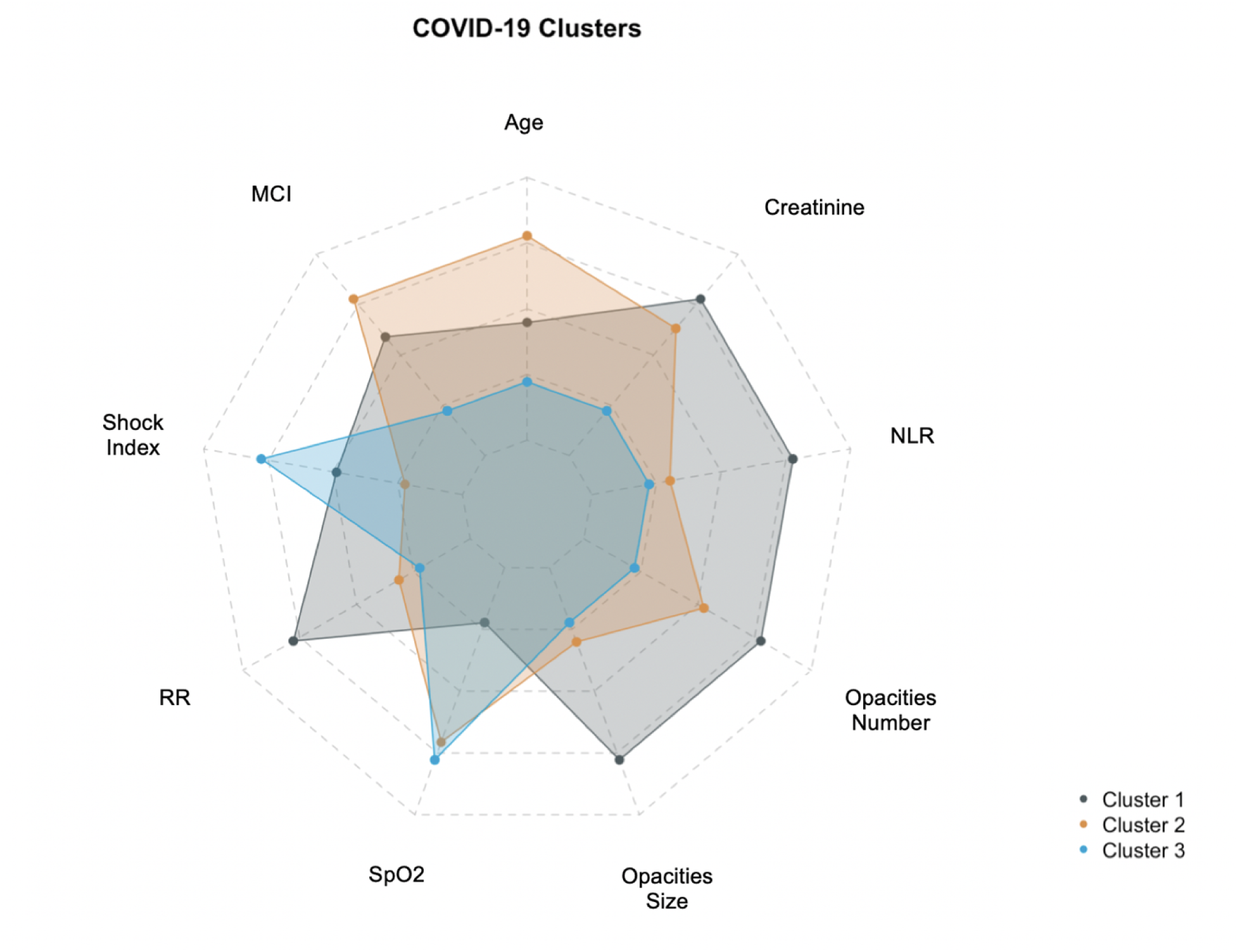
Radar Plot showing the distribution of clinical variables across clusters

**Figure 3B.**
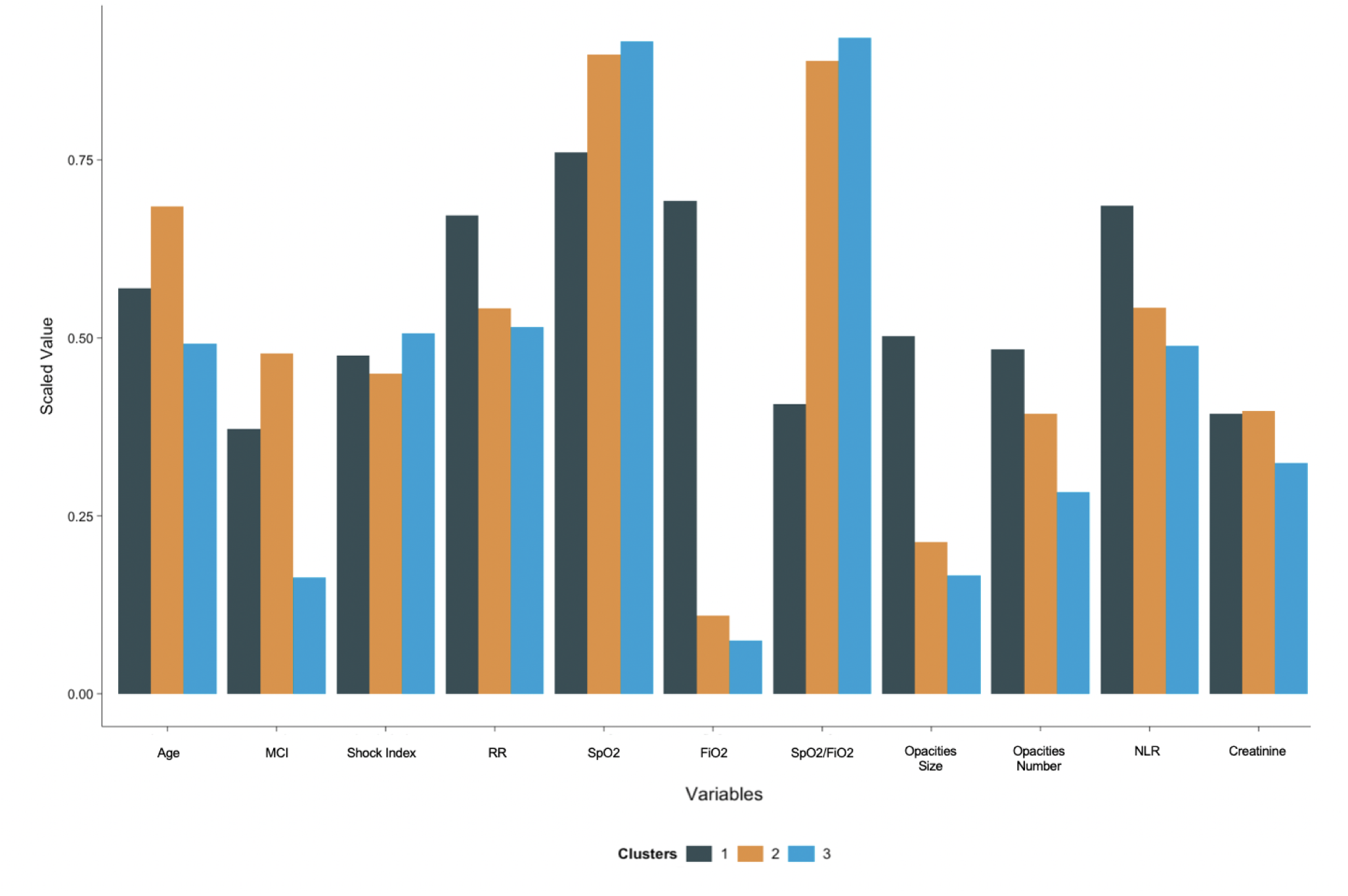
Bar Plot showing the distribution of clinical variables across clusters In both figures, the variables were scaled using normalization. Non-continuous variables underwent log-transformation if the skewness was > 0.5 and the respective mean of each scaled variable was plotted.

Cluster 1 (*n* = 61, 11%) represented the group of patients with the most severe presentation having the highest NLR (median 13.5), the highest rate of hypoxemia (median SpO_2_/FiO2 118) and the highest radiographic burden, with 91% of patients with at least two pulmonary opacities and a median total opacities size of 20% (ratio of opacities area/total CXR area).

Cluster 2 (*n* = 211, 39%) and Cluster 3 (*n* = 275, 50%) were similar regarding the respiratory, hematologic and hemodynamic subdomains. They both presented milder clinical manifestations than Cluster 1, but both phenotypes differed in terms of demographics. Cluster 2 represented the oldest cohort (mean 75.4 years) and Cluster 3 represented the youngest cohort (mean 59.8 years). They also differed by MCI, as Cluster 2 represented the group with the highest proportion of comorbidities, with a mean MCI of 4.30, compared to 1.47 for Cluster 3. Accordingly, Cluster 2 included patients with high proportions of concurrent medication on admission: 68% took anti-hypertensive agents, 63% hypolipemiant agents, 56.9% hypoglycemic agents, 53.6% antiplatelets agents and 37.4% bronchodilator agents.

### Phenotypes and clinical outcomes

Among the 547 patients in our study cohort, 436 patients were eligible for ICU admission and MV as deemed by their POLST form, and were thus analyzed for clinical outcomes (see **Fig 1**).

The cumulative mortality risk was significantly different across clusters (log-rank test, *p* = 0.01). The 30-day mortality risks were respectively 39% (21-53%, 95 CI), 33% (22-43%, 95 CI) and 20% (10-28%, 95 CI) for clusters 1, 2 and 3. The difference in cumulative ICU admission risk and cumulative mechanical ventilation risk were also statistically significant across all clusters (log-rank test respectively, *p*< 0.0001 and *p*< 0.0001). More precisely, the 7-day ICU admission risk was 74% (60-83%, 95% CI) for Cluster 1, 28% for Cluster 2 (20-35%, 95% CI) and 21% (15-26%, 95% CI) for Cluster 3. The 7-day mechanical ventilation risk was 58% (43-70%, 95% CI) for Cluster 1, 14% (8-19%, 95% CI) for Cluster 2 and 11% (6-15%, 95% CI) for Cluster 3 (**see Fig 4)**.

**Figure 4.**
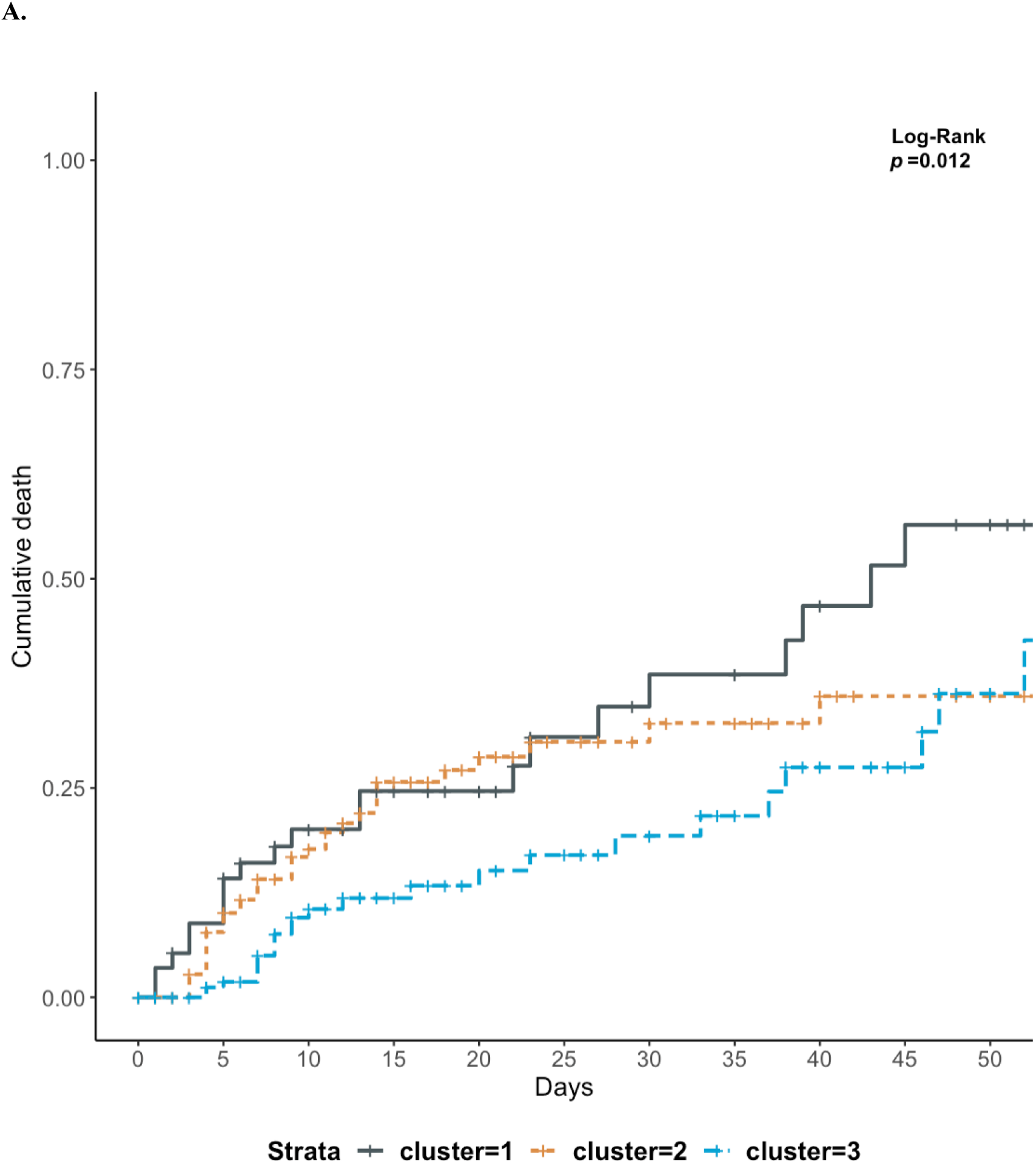
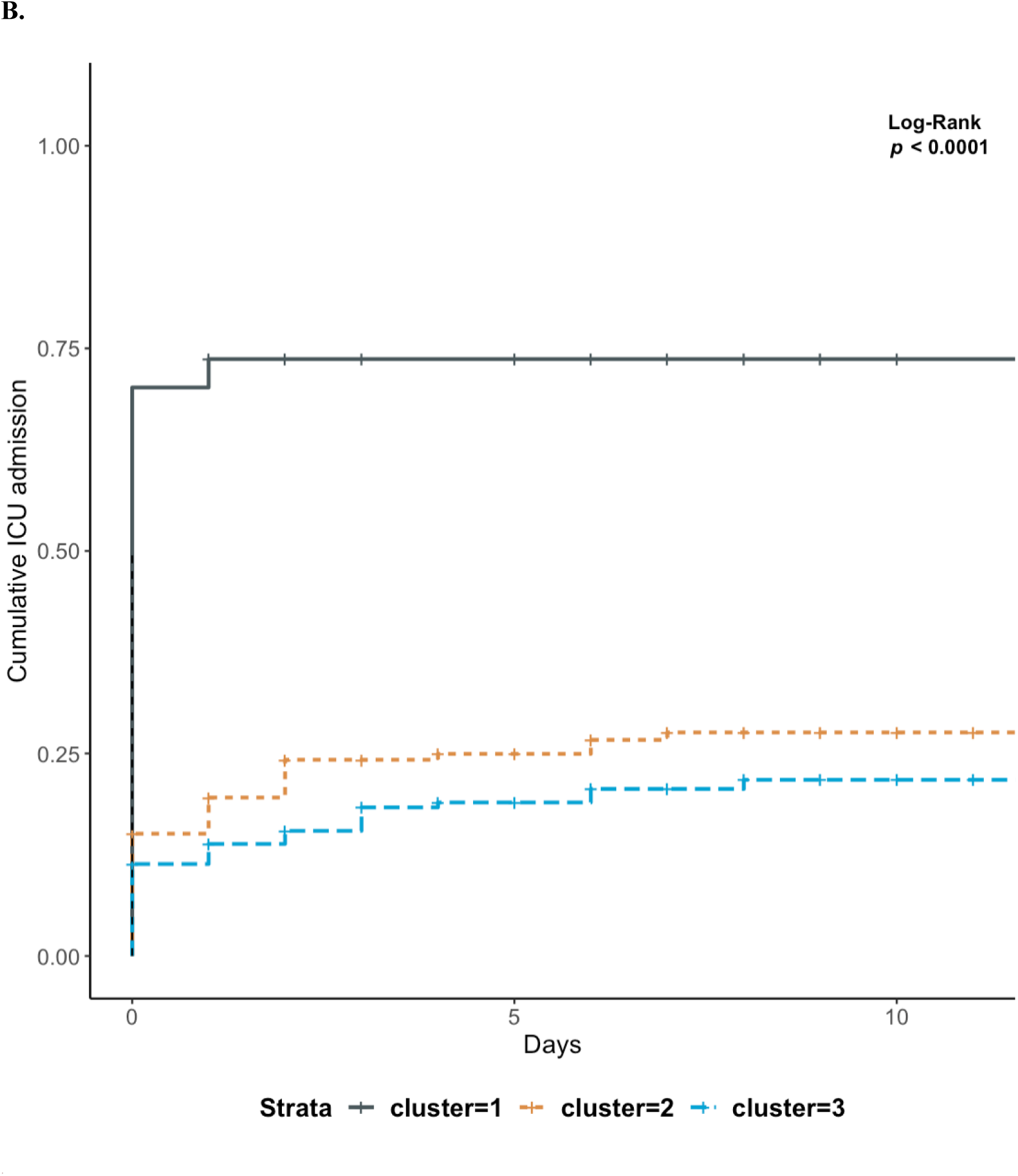
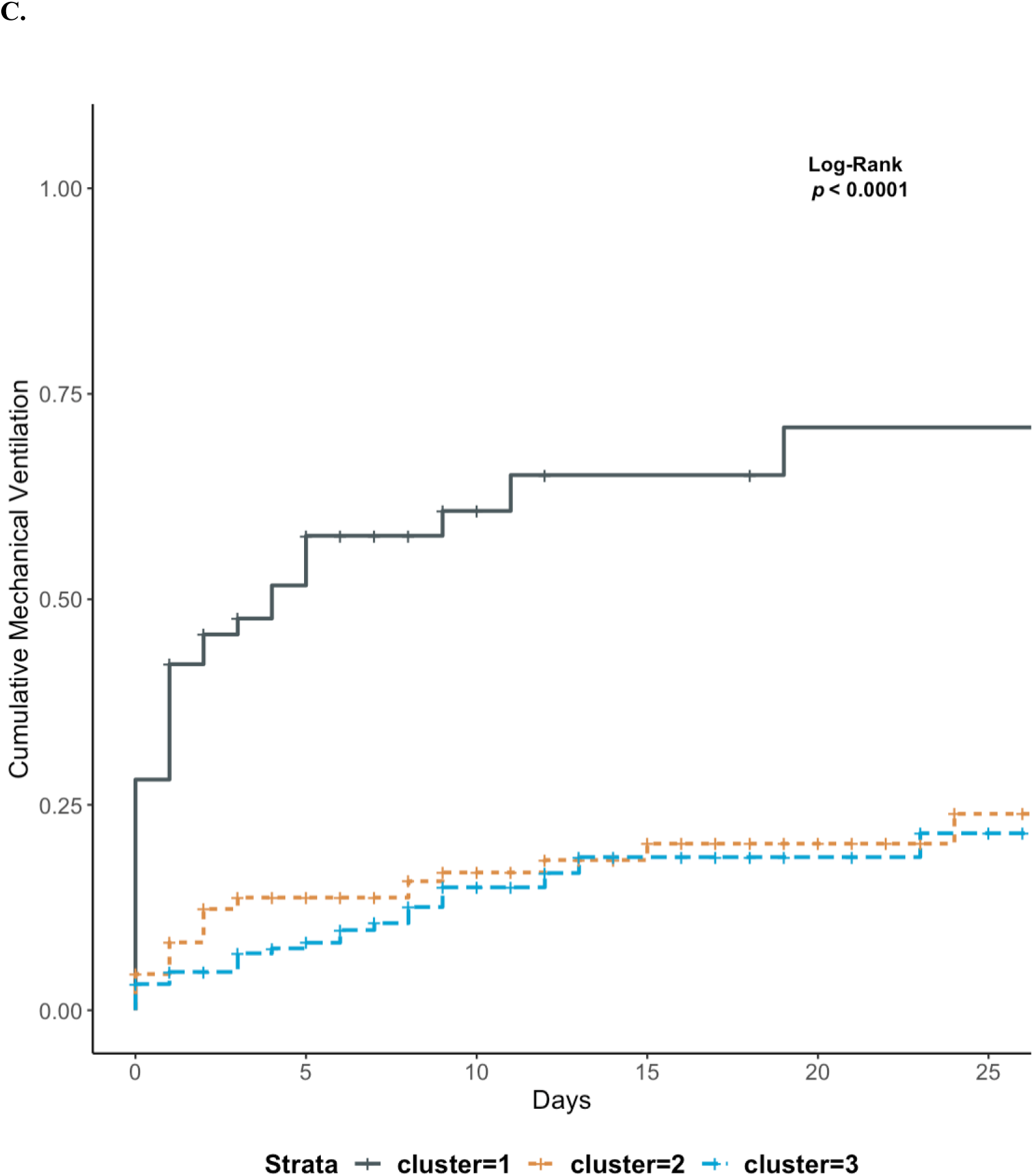
Kaplan-Meier curves for clinical outcomes **(a)** death, **(b)** ICU admission, and **(c)** mechanical ventilation stratified by phenotypes.

### Phenotypes Interpretability and Assignment

We developed a simple decision tree that allows the assignment of patients to their respective clinical phenotypes using the rules shown in **Fig 5**. Using only four variables (age, MCI, SpO_2_/FiO_2_, opacities size) and by following between three and five steps, one can assign a patient to one of the three phenotypes with an accuracy varying between 63 and 100% according to the given path.

**Figure 5.**
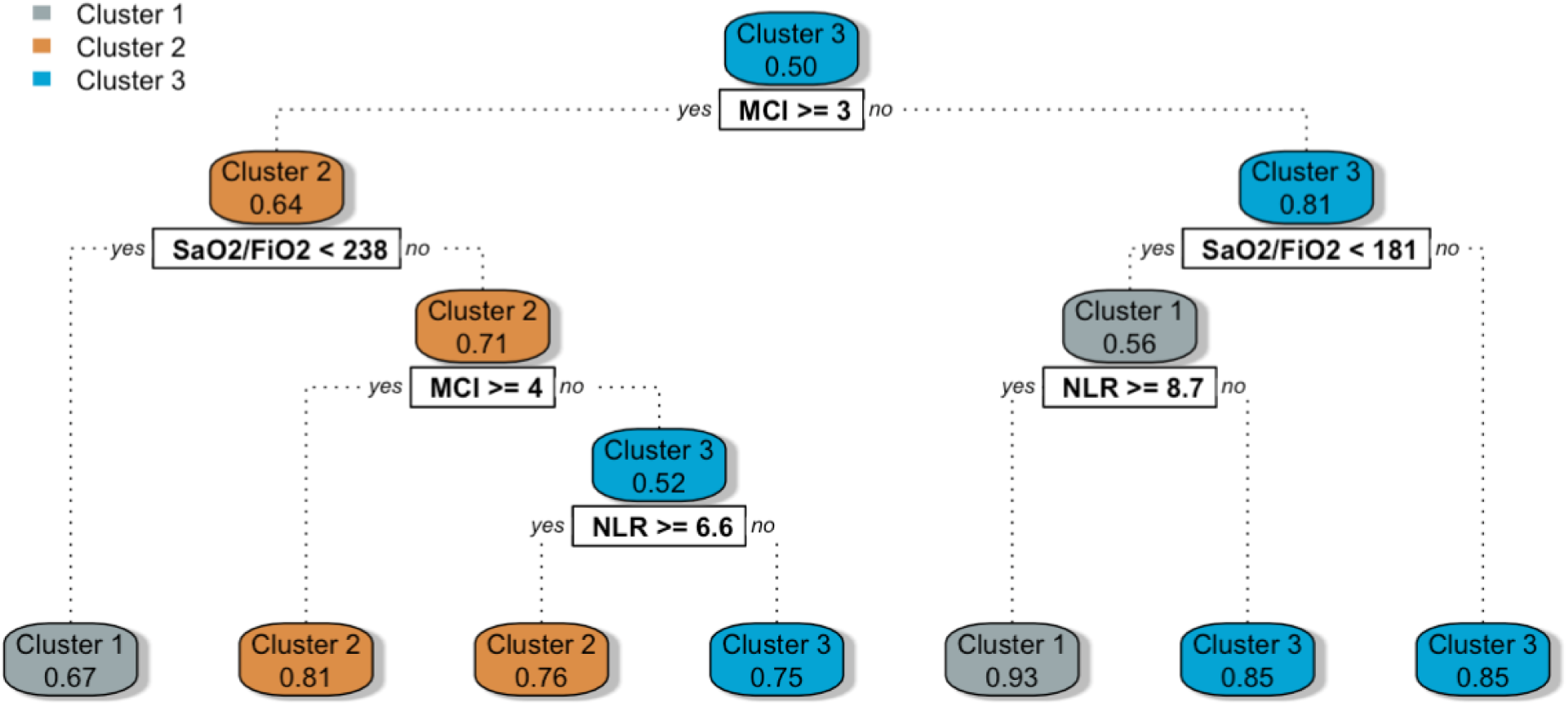
Clusters assignment through decision tree-based rules This graphs represents the classification rules obtained after training a CART decision tree algorithm with the phenotypes as outcomes. The rules obtained could be used at the bedside to determine the phenotype of a given individual following simple steps. The rules are located in the white squared boxes and each colored node box displays the probability of the predicted class.

### Phenotypes Robustness: sensitivity analyses

When comparing the clustering results with and without the inclusion of imaging data, the adjusted rand index was 0.55 indicating that the similarity of the two clusterings was moderate. The average silhouette width remained unchanged (0.34 with imaging; 0.33 without imaging), indicating that both approaches yielded homogeneous clusters.

We then further characterized the individuals who underwent phenotypic reclassification. 16 percent of patients (*n* = 87) underwent reclassification (see **Fig 6**), meaning that their clusters assignment changed after the removal of imaging data. The highest proportion of reclassified observations came from Cluster 1 as 32 percent of observations (*n* = 20) were reclassified to Cluster 2 or 3 after the removal of imaging data. When analyzing the clinical outcomes of those patients, we noted that 11 out of 20 patients died (55%). Conversely, we noted that eight patients who were initially assigned to Cluster 2 or 3, were reassigned to Cluster 1 (the most severe phenotype) after the removal of imaging data from the algorithm. All of those eight patients survived.

**Figure 6.**
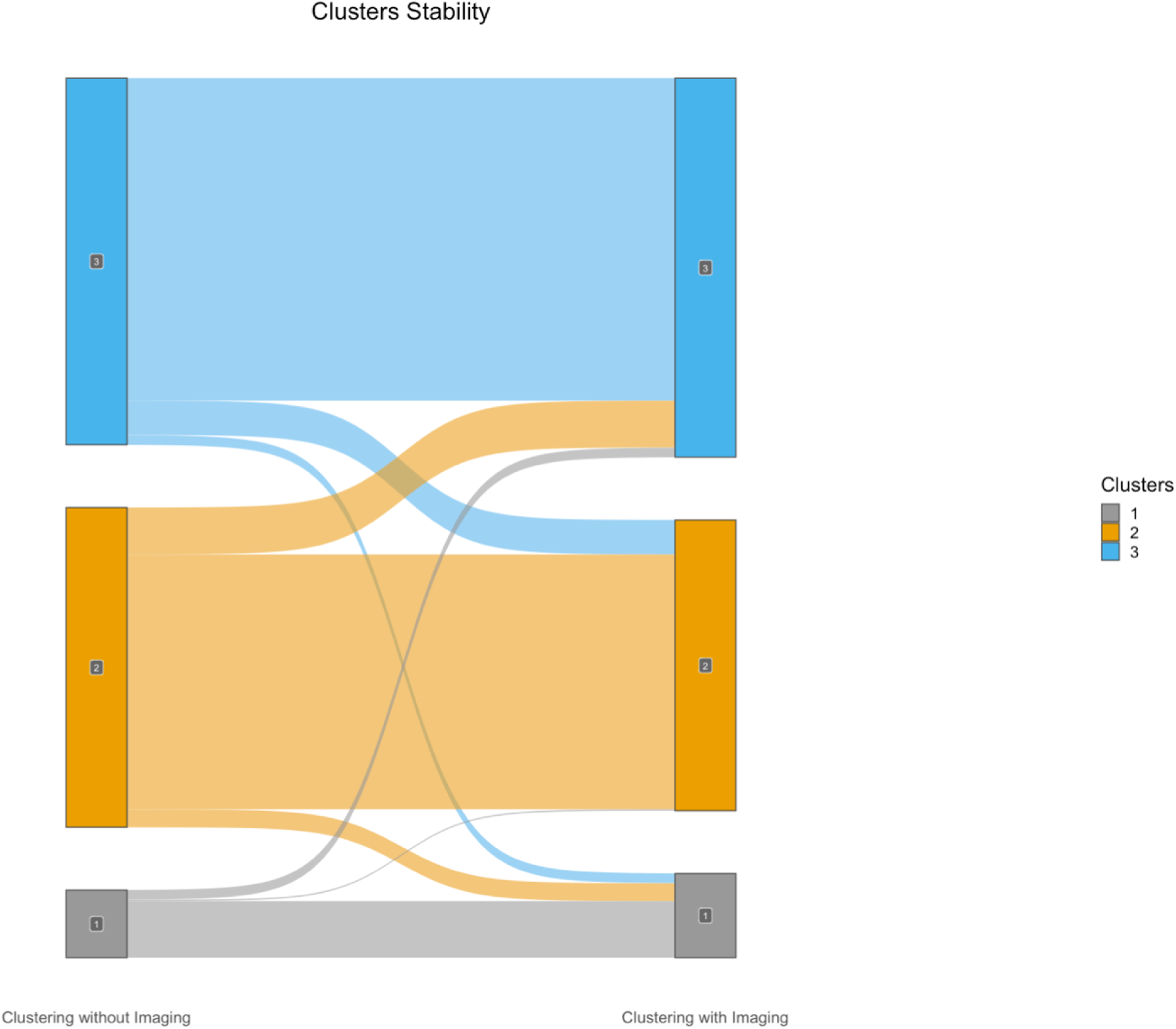
Sankey diagram assessing clustering stability with and without imaging data This plot shows the distribution of the patients that underwent reclassification after the removal of imaging data in the clustering algorithm. 87 observations (16%) were reclassified when ignoring imaging data. Although the clusters are relatively stable, a disproportionate number of reclassified observations originate from Cluster 1, as 20 observations (32%) were reclassified after the removal of imaging. As Cluster 1 is the most severe phenotype, the potential impact of this reclassification is not without consequences.

## Discussion

We identified three clinical phenotypes with distinct clinical characteristics and outcomes using multimodal clinical data in patients admitted with COVID-19. The three phenotypes can be summarized as follows: severely hypoxemic with high radiological burden irrespective of age (Cluster 1), mildly hypoxemic with either a high comorbidity index or old age (Cluster 2), and mildly hypoxemic with a low comorbidity index (Cluster 3).

The identified phenotypes were clinically relevant as they were associated with distinct clinical outcomes. Cluster 1 included patients with the most severe presentation and was thus unsurprisingly the phenotype with the highest mortality and morbidity risk. Cluster 2 and Cluster 3 represented patients with similar milder clinical presentations but with distinct comorbidities profiles. Patients in Cluster 2 had a higher comorbidity index (4.30 vs 1.47) and the 30-day mortality was higher when compared to Cluster 3 (32% vs. 21%). Being able to distinguish phenotypes with appearing similar features, but different outcomes, is clinically meaningful. Those represent patients currently treated identically, but who might benefit from a distinct and targeted treatment approach.

When comparing our results with previous work (33–40), the number of clusters obtained is consistent, as all have identified three phenotypes. However, all cited studies have reported phenotypes suggesting a linear relationship with age and disease severity. This drastically differs from our findings, in which the most severe phenotype (Cluster 1) does not represent the oldest group. In our opinion, our phenotypes reflect the complexity of the distribution of patients with COVID-19 in which age is not the sole determinant of severity. Further research is needed to understand the virological and immunological factors associated with severe infection in this phenotype.

Despite having considered more than 30 variables in our clustering algorithm, only four subdomains were central in establishing the phenotypes: demographics, hematologic features, respiratory features, and imaging data. Socio-demographics, comorbidities (41) and hypoxemia’s (42) impact on the clinical course of patients with COVID-19 have been well documented, and their relative importance in our clustering effort was thus expected. Furthermore, neutrophil-to-lymphocyte ratio (NLR), previously identified as an independent risk predictor for mortality in COVID-19 (43), was accordingly higher in Cluster 1. On the other hand, mean platelet volume (MPV) was not found to significantly impact clustering results despite being associated with severe forms of the disease (44).

Our study emphasizes the importance of imaging data in COVID-19 related clustering efforts. Through our sensitivity analysis, we have shown that the incorporation of CXR enhanced the phenotypes’ clinical value. Even in the absence of ground truth in unsupervised machine learning, we showed that dismissing imaging data reduced the clinical accuracy of our clustering algorithm. Eight patients initially assigned to Cluster 2 or 3 were reclassified to the more severe Cluster 1 after the removal of imaging data from the algorithm. We deemed the initial assignment to the less severe Cluster 2 or 3 as the accurate one, given all eight of those patients survived. Likewise, 20 patients initially assigned to Cluster 1 were reclassified to Cluster 2 or 3 after the removal of imaging data. We deemed the initial assignment to the severe Cluster 1 as appropriate, because the mortality rate of those 20 patients (55%) exceeded those of Cluster 2 and 3 (respectively 24% and 14%). The use of imaging helped properly classify outlier patients: those who had a lower or higher radiological burden than the majority of the patients in their respective phenotypic group (see **S3 Figure, S4 Figure**).

To our knowledge, opacity size has not previously been used in other COVID-19 phenotyping efforts. Instead, the number of opacities has been used as a proxy variable in only one other paper (33). The number of opacities is generally more accessible as it can directly be extracted from CXR reports, and does not demand manual annotation of medical images. However, studies have shown that even though those two variables provide overlapping information, they are not interchangeable and their respective value differs when it comes to predicting survival and the need for respiratory support in patients with COVID-19 (45). Furthermore, automation of chest X-ray opacities annotation is increasingly feasible with publicly available deep-learning models harmonizing the process (17). We opted for manual annotation in this paper, awaiting further validation of those tools in the COVID-19 population. Nonetheless, chest X-ray annotation should not be viewed as a rate-limiting process. On the contrary, it should be encouraged in healthcare machine learning as it allows uniformizing the inputs during training and upon model deployment (46).

Our study highlights the feasibility and the importance of agnostic approaches to disease phenotyping with no a priori information about patient outcome.

At the bedside, clinical phenotypes help unbiasedly categorize patients. Previous studies have shown that individual risk factors taken alone are insufficient to properly stratify patients with COVID-19 (45). Phenotypes thus offer a simple, and yet holistic means to describing patients with COVID-19 while incorporating both clinical presentation and morbidity risk.

In clinical trials, phenotypes could help harmonizing enrolled participants and facilitate the identification of patients’ subgroups benefiting from a given therapy. Recent clinical trials revealed that distinct clinical presentations mandate distinct treatments, with some therapies showing benefits only in patients with severe disease (47,48). The various inclusion criteria of these trials have made it difficult for clinicians to clearly identify patients who would benefit the most from those novel interventions. The controversy regarding the benefits of anticoagulation in critically and non-critically ill patients is a testimony of this observation (49). Using standardized phenotypes could remove the ambiguous nature of patients subgrouping in future trials and facilitate the comparison of results across trials.

Our study also highlights the clinical usability of clustering-based phenotypes (50). Variables needed to assign patients to phenotypes are readily available at the point-of-care, and cluster assignment can be done following three and five simple steps. Our decision-tree model for cluster assignment would need to be externally validated before being used in the clinical setting, but it nevertheless remains helpful to understand how the three phenotypes differ and how applicable such phenotypes would be in the clinical setting. Moreover, the probabilistic nature of the algorithm allows for direct quantification of the uncertainty, which is an important factor for clinical usability of machine learning algorithms (51). We restricted our clustering effort to the data available within the first 24 hours of admission to ease generalizability and to emulate the timing of triage decision that is often made early upon admission. To our knowledge, characterizing the evolution of phenotypes over time has not been studied and would mandate further investigation.

We recognize that clinical phenotypes do not offer a comprehensive explanation model for the observed disease heterogeneity (52). However, they lay the groundwork for understanding COVID-19 pathobiology. Studies linking biobank data to clinical phenotypes allow to capture the taxonomic complexity of the disease and describe how phenotypes differ in terms of pathogenic mechanisms (53).

Our study presents some limitations. Multiple variables could not be included because they were either not captured in our electronic health record (e.g., time from onset of symptoms, mechanical ventilation parameters, in-hospital complications) or excluded from our study because of missingness. However, missing values are common in clinical practice and investigating risk stratification while considering the inherent characteristics real-world data is of importance at the bedside (54). In addition, this enhances the applicability of our phenotypes, as they are only based on the most common variables available for patients admitted with COVID-19 (55). This differs from studies that have included flux cytometry and CD4+/CD8+ count in their algorithm (34). Besides, those *omitted* variables do not seem to have had significant impact on our results as the three clusters obtained were consistent in numbers with previous work (33–39).

Additionally, our study included patients admitted between January 1, 2020, and January 31, 2021, being before the approval of the majority of targeted therapies against COVID-19 or vaccination. We therefore did not assess the effect of vaccination, treatments and the type of variant on phenotypes. Accordingly, this put our algorithm at risk for temporal dataset shift (56) and calibrating our clustering algorithm will be necessary before exploiting it in the clinical setting. Finally, because race-based data is not recorded in the Quebec healthcare system (57), we could not proceed to sensitivity analysis according to race. For that very same reason, we acknowledge that our work could be subject to algorithmic bias as evidence has shown racial disparities in clinical outcomes of patients with COVID-19 (58).

## Conclusion

We developed a multidimensional phenotypic analysis of patients with COVID-19 and identified three distinct phenotypes, with one specifically associated with worse clinical outcomes. Our study supports the feasibility of using real-world clinical data to conduct unsupervised phenotypic clustering. External validation and further research are needed to determine how phenotypes could impact clinical trials design and phenotypic-guided treatment in clinical practice.

## Data Availability

The entire code excluding the dataset is publicly available on GitHub (https://github.com/CODA-19/models/tree/master/phenotyper) The data that support the findings of this study are available on request from the corresponding author, MC.

## Abbreviations

APN: average proportion of non-overlap
AD: average distance
ADM: average distance between means
CART: classification and regression tree
CCI: Charlson Comorbidity Index
CXR: chest radiographs
FAMD: factor analysis of mixed data
FOM: figure of merit
ICU: intensive care unit
MCI: Medicines Comorbidity Index
MV: mechanical ventilation
NLR: neutrophil-to-lymphocyte ratio
PAM: partition around medoids
PCR: polymerase chain reaction
POLST: Physician Orders for Life-Sustaining Treatment
VIA: variable importance analysis

## Acknowledgments

M.C. is supported by a Fonds de Recherche Québec Santé (FRQS) Clinical Research scholarship. M.D. is supported by a FRQS Clinical Research scholarship. A.T. is supported by a FRQS Clinical Research Scholarship and a Fondation de l’Association des Radiologistes du Québec (FARQ) Clinical Research Scholarship. This work was possible with the contribution of the CHUM Center for Integration and Analysis of Medical Data (CITADEL).

This work was partly funded by a Quebec Bio-Imaging Network research grant. The funders had no role in study design, data collection and analysis, decision to publish, or preparation of the manuscript.

## Reproducibility

The entire code excluding the dataset is publicly available on GitHub (https://github.com/CODA-19/models/tree/master/phenotyper).

The data that support the findings of this study are available on request from the corresponding author, MC.

## Supporting information

S1 text (Supplementary Methods)

S1-S3 Tables

S1-S4 Figures

## References

1. The Lancet Rheumatology. High-stakes heterogeneity in COVID-19. Lancet Rheumatol. 2020 Oct;2(10):e577.

2. Wojczynski MK, Tiwari HK. Definition of Phenotype. Genetic dissection of complex traits. Elsevier; 2008. p. 75–105.

3. Gattinoni L, Coppola S, Cressoni M, Busana M, Rossi S, Chiumello D. COVID-19 Does Not Lead to a “Typical” Acute Respiratory Distress Syndrome. Am J Respir Crit Care Med. 2020 May 15;201(10):1299–300.

4. Gattinoni L, Chiumello D, Caironi P, Busana M, Romitti F, Brazzi L, et al. COVID-19 pneumonia: different respiratory treatments for different phenotypes? Intensive Care Med. 2020 Jun;46(6):1099–102.

5. Spiezia L, Boscolo A, Poletto F, Cerruti L, Tiberio I, Campello E, et al. COVID-19-Related Severe Hypercoagulability in Patients Admitted to Intensive Care Unit for Acute Respiratory Failure. Thromb Haemost. 2020 Jun;120(6):998–1000.

6. Spyropoulos AC, Goldin M, Giannis D, Diab W, Wang J, Khanijo S, et al. Efficacy and Safety of Therapeutic-Dose Heparin vs Standard Prophylactic or Intermediate-Dose Heparins for Thromboprophylaxis in High-risk Hospitalized Patients With COVID-19: The HEP-COVID Randomized Clinical Trial. JAMA Intern Med. 2021 Dec 1;181(12):1612–20.

7. Tang N, Li D, Wang X, Sun Z. Abnormal coagulation parameters are associated with poor prognosis in patients with novel coronavirus pneumonia. J Thromb Haemost JTH. 18(4):844–7.

8. Gattinoni L, Camporota L, Marini JJ. COVID-19 phenotypes: leading or misleading? Eur Respir J. 2020 Aug 27;56(2).

9. Jain A, Doyle DJ. Stages or phenotypes? A critical look at COVID-19 pathophysiology. Intensive Care Med. 2020 Jul;46(7):1494–5.

10. Bzdok D, Altman N, Krzywinski M. Statistics versus machine learning. Nat Methods. 2018 Apr 3;15(4):233–4.

11. Siroux V, Basagaña X, Boudier A, Pin I, Garcia-Aymerich J, Vesin A, et al. Identifying adult asthma phenotypes using a clustering approach. Eur Respir J. 2011 Aug;38(2):310–7.

12. Deliu M, Sperrin M, Belgrave D, Custovic A. Identification of asthma subtypes using clustering methodologies. Pulm Ther. 2016 Jun 22;2:19–41.

13. Castaldi PJ, Benet M, Petersen H, Rafaels N, Finigan J, Paoletti M, et al. Do COPD subtypes really exist? COPD heterogeneity and clustering in 10 independent cohorts. Thorax. 2017 Nov;72(11):998–1006.

14. Seymour CW, Kennedy JN, Wang S, Chang C-CH, Elliott CF, Xu Z, et al. Derivation, validation, and potential treatment implications of novel clinical phenotypes for sepsis. JAMA. 2019 May 28;321(20):2003–17.

15. Horne E, Tibble H, Sheikh A, Tsanas A. Challenges of clustering multimodal clinical data: review of applications in asthma subtyping. JMIR Med Inform. 2020 May 28;8(5):e16452.

16. GitHub - izolot/BBox-Label-Tool-Python3.x: A simple tool for labeling object bounding boxes in images Python 3.X [Internet]. [cited 2021 Oct 6]. Available from: https://github.com/izolot/BBox-Label-Tool-Python3.x

17. Pan I, Cadrin-Chênevert A, Cheng PM. Tackling the radiological society of north america pneumonia detection challenge. AJR Am J Roentgenol. 2019 Sep;213(3):568–74.

18. Shih G, Wu CC, Halabi SS, Kohli MD, Prevedello LM, Cook TS, et al. Augmenting the National Institutes of Health Chest Radiograph Dataset with Expert Annotations of Possible Pneumonia. Radiol Artif Intell. 2019 Jan 30;1(1):e180041.

19. Tang F, Ishwaran H. Random forest missing data algorithms. Stat Anal Data Min. 2017 Dec;10(6):363–77.

20. Duy Le T, Beuran R, Tan Y. Comparison of the most influential missing data imputation algorithms for healthcare. 2018 10th International Conference on Knowledge and Systems Engineering (KSE). IEEE; 2018. p. 247–51.

21. Yang A-P, Liu J-P, Tao W-Q, Li H-M. The diagnostic and predictive role of NLR, d-NLR and PLR in COVID-19 patients. Int Immunopharmacol. 2020 Jul;84:106504.

22. Catoire P, Tellier E, de la Rivière C, Beauvieux M-C, Valdenaire G, Galinski M, et al. Assessment of the SpO2/FiO2 ratio as a tool for hypoxemia screening in the emergency department. Am J Emerg Med. 2021 Jun;44:116–20.

23. Doğanay F, Elkonca F, Seyhan AU, Yilmaz E, Batirel A, Ak R. Shock index as a predictor of mortality among the Covid-19 patients. Am J Emerg Med. 2021 Feb;40:106–9.

24. Narayan SW, Nishtala PS. Development and validation of a Medicines Comorbidity Index for older people. Eur J Clin Pharmacol. 2017 Dec;73(12):1665–72.

25. Alelyani S, Tang J, Liu H. Feature selection for clustering: A review. In: Aggarwal CC, Reddy CK, editors. Data clustering: algorithms and applications. Chapman and Hall/CRC; 2018. p. 29–60.

26. Nestor B, McDermott MBA, Chauhan G, Naumann T, Hughes MC, Goldenberg A, et al. Rethinking clinical prediction: Why machine learning must consider year of care and feature aggregation. arXiv. 2018;

27. Ding C, He X. K-means clustering via principal component analysis. Proceedings of the twenty-first international conference on Machine learning. 2004;29.

28. Sekula M, Datta S, Datta S. optCluster: An R Package for Determining the Optimal Clustering Algorithm. Bioinformation. 2017 Mar 31;13(3):101–3.

29. Santos JM, Embrechts M. On the use of the adjusted rand index as a metric for evaluating supervised classification. In: Alippi C, Polycarpou M, Panayiotou C, Ellinas G, editors. Artificial neural networks – ICANN 2009. Berlin, Heidelberg: Springer Berlin Heidelberg; 2009. p. 175–84.

30. Lewis RJ. An introduction to classification and regression tree (CART) analysis. Annual meeting of the society for academic emergency medicine in San Francisco, California. 2000;14.

31. Wei P, Lu Z, Song J. Variable importance analysis: A comprehensive review. Reliability Engineering & System Safety. 2015 Oct;142:399–432.

32. Asch DA, Sheils NE, Islam MN, Chen Y, Werner RM, Buresh J, et al. Variation in US Hospital Mortality Rates for Patients Admitted With COVID-19 During the First 6 Months of the Pandemic. JAMA Intern Med. 2021 Apr 1;181(4):471–8.

33. Gutiérrez-Gutiérrez B, Del Toro MD, Borobia AM, Carcas A, Jarrín I, Yllescas M, et al. Identification and validation of clinical phenotypes with prognostic implications in patients admitted to hospital with COVID-19: a multicentre cohort study. Lancet Infect Dis. 2021 Jun;21(6):783–92.

34. Ye W, Lu W, Tang Y, Chen G, Li X, Ji C, et al. Identification of COVID-19 Clinical Phenotypes by Principal Component Analysis-Based Cluster Analysis. Front Med (Lausanne). 2020 Nov 12;7:570614.

35. Lascarrou J-B, Gaultier A, Soumagne T, Serck N, Sauneuf B, Piagnerelli M, et al. Identifying Clinical Phenotypes in Moderate to Severe Acute Respiratory Distress Syndrome Related to COVID-19: The COVADIS Study. Front Med (Lausanne). 2021 Mar 11;8:632933.

36. Rubio-Rivas M, Corbella X, Mora-Luján JM, Loureiro-Amigo J, López Sampalo A, Yera Bergua C, et al. Predicting Clinical Outcome with Phenotypic Clusters in COVID-19 Pneumonia: An Analysis of 12,066 Hospitalized Patients from the Spanish Registry SEMI-COVID-19. J Clin Med. 2020 Oct 29;9(11).

37. Azoulay E, Zafrani L, Mirouse A, Lengliné E, Darmon M, Chevret S. Clinical phenotypes of critically ill COVID-19 patients. Intensive Care Med. 2020 Aug;46(8):1651–2.

38. Lusczek ER, Ingraham NE, Karam BS, Proper J, Siegel L, Helgeson ES, et al. Characterizing COVID-19 clinical phenotypes and associated comorbidities and complication profiles. PLoS One. 2021 Mar 31;16(3):e0248956.

39. Rodríguez A, Ruiz-Botella M, Martín-Loeches I, Jimenez Herrera M, Solé-Violan J, Gómez J, et al. Deploying unsupervised clustering analysis to derive clinical phenotypes and risk factors associated with mortality risk in 2022 critically ill patients with COVID-19 in Spain. Crit Care. 2021 Feb 15;25(1):63.

40. Batah SS, Benatti MN, Siyuan L, Telini WM, Barboza JO, Menezes MB, et al. COVID-19 bimodal clinical and pathological phenotypes. Clin Transl Med. 2022 Jan;12(1):e648.

41. Bhattacharyya A, Seth A, Srivast N, Imeokparia M, Rai S. Coronavirus (COVID-19): A Systematic Review and Meta-analysis to Evaluate the Significance of Demographics and Comorbidities. Res Sq. 2021 Jan 18;

42. Xie J, Covassin N, Fan Z, Singh P, Gao W, Li G, et al. Association Between Hypoxemia and Mortality in Patients With COVID-19. Mayo Clin Proc. 2020 Jun;95(6):1138–47.

43. Huang I, Pranata R. Lymphopenia in severe coronavirus disease-2019 (COVID-19): systematic review and meta-analysis. J Intensive Care. 2020 May 24;8:36.

44. Lippi G, Henry BM, Favaloro EJ. Mean Platelet Volume Predicts Severe COVID-19 Illness. Semin Thromb Hemost. 2021 Jun;47(4):456–9.

45. Balbi M, Caroli A, Corsi A, Milanese G, Surace A, Di Marco F, et al. Chest X-ray for predicting mortality and the need for ventilatory support in COVID-19 patients presenting to the emergency department. Eur Radiol. 2021 Apr;31(4):1999–2012.

46. Zunair H, Rahman A, Mohammed N, Cohen JP. Uniformizing Techniques to Process CT Scans with 3D CNNs for Tuberculosis Prediction. In: Rekik I, Adeli E, Park SH, Valdés Hernández M del C, editors. Predictive Intelligence in Medicine: Third International Workshop, PRIME 2020, Held in Conjunction with MICCAI 2020, Lima, Peru, October 8, 2020, Proceedings. Cham: Springer International Publishing; 2020. p. 156–68.

47. Robba C, Battaglini D, Ball L, Patroniti N, Loconte M, Brunetti I, et al. Distinct phenotypes require distinct respiratory management strategies in severe COVID-19. Respir Physiol Neurobiol. 2020 Aug;279:103455.

48. Health OW. Therapeutics and COVID-19: living guideline, 14 January 2022. Therapeutics and COVID-19: living guideline, 14 January 2022. 2022;

49. Jorda A, Siller-Matula JM, Zeitlinger M, Jilma B, Gelbenegger G. Anticoagulant Treatment Regimens in Patients With Covid-19: A Meta-Analysis. Clin Pharmacol Ther. 2022 Mar;111(3):614–23.

50. Cutillo CM, Sharma KR, Foschini L, Kundu S, Mackintosh M, Mandl KD, et al. Machine intelligence in healthcare-perspectives on trustworthiness, explainability, usability, and transparency. npj Digital Med. 2020 Mar 26;3:47.

51. Chen IY, Joshi S, Ghassemi M, Ranganath R. Probabilistic machine learning for healthcare. Annu Rev Biomed Data Sci. 2021 Jul 20;4:393–415.

52. DeMerle K, Angus DC, Seymour CW. Precision Medicine for COVID-19: Phenotype Anarchy or Promise Realized? JAMA. 2021 May 25;325(20):2041–2.

53. Osuchowski MF, Winkler MS, Skirecki T, Cajander S, Shankar-Hari M, Lachmann G, et al. The COVID-19 puzzle: deciphering pathophysiology and phenotypes of a new disease entity. Lancet Respir Med. 2021 Jun;9(6):622–42.

54. Yu M, Tang A, Brown K, Bouchakri R, St-Onge P, Wu S, et al. Integrating artificial intelligence in bedside care for covid-19 and future pandemics. BMJ. 2021 Dec 31;375:e068197.

55. Brat GA, Weber GM, Gehlenborg N, Avillach P, Palmer NP, Chiovato L, et al. International electronic health record-derived COVID-19 clinical course profiles: the 4CE consortium. npj Digital Med. 2020 Aug 19;3:109.

56. Guo LL, Pfohl SR, Fries J, Posada J, Fleming SL, Aftandilian C, et al. Systematic review of approaches to preserve machine learning performance in the presence of temporal dataset shift in clinical medicine. Appl Clin Inform. 2021 Aug;12(4):808–15.

57. Jean-pierre J, Collins T. The effect of COVID-19 on Black communities in Quebec.

58. Kopel J, Perisetti A, Roghani A, Aziz M, Gajendran M, Goyal H. Racial and Gender-Based Differences in COVID-19. Front Public Health. 2020 Jul 28;8:418.

59. Bodenreider O, Peters LB, Nguyen T. RxClass-Navigating between Drug Classes and RxNorm Drugs. ICBO. 2014;106.

60. Gutiérrez-Sacristán A, Bravo À, Giannoula A, Mayer MA, Sanz F, Furlong LI. comoRbidity: an R package for the systematic analysis of disease comorbidities. Bioinformatics. 2018 Sep 15;34(18):3228–30.

61. Austin SR, Wong Y-N, Uzzo RG, Beck JR, Egleston BL. Why Summary Comorbidity Measures Such As the Charlson Comorbidity Index and Elixhauser Score Work. Med Care. 2015 Sep;53(9):e65–72.

62. Budiaji W, Leisch F. Simple K-Medoids Partitioning Algorithm for Mixed Variable Data. Algorithms. 2019 Aug 24;12(9):177.

63. Arora S, Hu W, Kothari PK. An Analysis of the t-SNE Algorithm for Data Visualization. 2018 Jul 3;

64. Murtagh F, Contreras P. Algorithms for hierarchical clustering: an overview. WIREs Data Mining Knowl Discov. 2012 Jan;2(1):86–97.

65. Gower JC, Warrens MJ. Similarity, dissimilarity, and distance, measures of. In: Balakrishnan N, Colton T, Everitt B, Piegorsch W, Ruggeri F, Teugels JL, editors. Wiley statsref: statistics reference online. Chichester, UK: John Wiley & Sons, Ltd; 2014. p. 1–11.

66. Pandit S, Gupta S. A comparative study on distance measuring approaches for clustering. IJORCS. 2011 Dec 30;2(1):29–31.

67. Nagpal A, Jatain A, Gaur D. Review based on data clustering algorithms. 2013 IEEE CONFERENCE ON INFORMATION AND COMMUNICATION TECHNOLOGIES. IEEE; 2013. p. 298–303.

68. Ahmad A, Khan SS. Survey of State-of-the-Art Mixed Data Clustering Algorithms. IEEE Access. 2019;7:31883–902.

69. Pihur V, Datta S, Datta S. RankAggreg, an R package for weighted rank aggregation. BMC Bioinformatics. 2009 Feb 19;10:62.

